# A Polygenic Score for Smoking Persistence Captures Differences in Cessation Age

**DOI:** 10.64898/2026.10.22.26366250

**Authors:** Ariya Chaloemtoem, Yoonhoo Chang, Louis Fox, Vikram R. Shaw, Jinyoung Byun, Tony Chen, Ricardo Cortez Cardoso Penha, Dana Hancock, Rayjean Hung, Eric O. Johnson, Alex T. Ramsey, Nancy L. Saccone, Gretchen Saunders, Haoyu Zhang, James McKay, Christopher Amos, Li-Shiun Chen, Laura J. Bierut

## Abstract

**Introduction:** Smoking cessation substantially reduces tobacco-related morbidity and mortality, with earlier cessation conferring greater health benefits. Polygenic scores (PGSs) for smoking persistence may provide insight into cessation timing across diverse populations. This study examined the association between a multi-ancestry PGS for smoking persistence and smoking cessation timing in a genetically diverse population.

**Methods:** This study included 139,129 current and former smokers with genetic data from the All of Us Research Program. A multi-ancestry PGS for smoking persistence was constructed using genome-wide association study summary statistics from the GWAS & Sequencing Consortium of Alcohol and Nicotine Use Phase 2 (GSCAN2). Smoking status and cessation timing were examined across the PGS distribution, with a focus on differences between the highest and lowest deciles. Median cessation ages and cumulative cessation probabilities were estimated, while Cox proportional hazards models compared cessation hazards across PGS groups overall and by genetic ancestry.

**Results:** Among 139,129 participants, the proportion of current smokers increased across the PGS distribution, from 35.5% in the lowest decile to 51.9% in the highest decile. Unadjusted median cessation ages were 38 and 50 years in the lowest and highest deciles, respectively (N = 125,047). After adjusting for sex, educational attainment, household income, cigarettes smoked per day, and genetic population structure, estimated median cessation ages were 40 years (95% CI: 40-40) and 48 years (95% CI: 47-50), respectively, representing an 8-year difference. Participants in the highest decile also had a lower cessation hazard than those in the lowest decile (HR = 0.67, 95% CI: 0.65-0.70), with associations observed across genetic ancestry groups, although estimates varied in precision.

**Conclusion:** A multi-ancestry PGS for smoking persistence was associated with substantial differences in cessation timing across genetically diverse populations. These findings suggest that genetic liability to smoking persistence may help characterize risk of delayed cessation and inform earlier, more targeted smoking cessation support.

## Introduction

Cigarette smoking remains a leading preventable cause of morbidity and mortality worldwide, contributing to a significant burden of diseases including lung cancer, chronic obstructive pulmonary disease (COPD), heart disease, and stroke.^1,2^ Although most adults who smoke report wanting to quit, successful cessation remains difficult, and many individuals require repeated quit attempts.^3^

Quitting smoking is beneficial at any age, but the timing of cessation has important implications for long-term health.^4,5^ Individuals who stop smoking before the age of 40 can avoid more than 90% of the excess mortality associated with continued smoking. While cessation even at older ages remains associated with substantial survival benefits, morbidity and mortality increase with later age of cessation.^5^ Benefits of cessation are also evident among individuals with established smoking-related disease, including improved survival following cancer diagnosis and reductions in subsequent cardiovascular disease risk.^6,7^ Thus, smoking cessation is not only a question of whether an individual eventually quits, but also when cessation occurs across the life course.

Individual differences in smoking cessation are partly attributable to genetic factors, with twin studies reporting heritability estimates of 51% to 58%.^8–10^ Large-scale genome-wide association studies (GWASs) have identified thousands of genetic variants associated with smoking behaviors, including smoking cessation.^11–13^ Although individual variants generally have small effects, their GWAS-estimated effects can be aggregated using polygenic scores (PGSs) to provide a single measure of genetic liability.^14^

In this study, we constructed a multi-ancestry PGS for smoking cessation and evaluated its associations with smoking cessation in the diverse All of Us (AoU) cohort. We examined associations with both cessation status and the timing of cessation across adulthood. We further characterized differences in smoking persistence and cessation timing across the PGS distribution, including individuals at the extremes of genetic liability, and assessed the consistency of these associations across genetically inferred ancestry groups. Together, these analyses aimed to extend our understanding of how polygenic liability relates to clinically important smoking cessation trajectories across adulthood in genetically diverse populations.

## Methods

### 1 Data Sources & Study Population

This study integrated genetic and phenotypic data from three sources to construct, calibrate, and evaluate a polygenic score for smoking cessation (Supplementary Figure S1).

#### 1.1 GSCAN2 Smoking Cessation GWAS Summary Statistics

Ancestry-specific summary statistics for smoking cessation (SmkCes) from the GWAS and Sequencing Consortium of Alcohol and Nicotine Use Phase 2 (GSCAN2),^13^ excluding 23andMe participants due to data restrictions, included 509,190 individuals of African (AFR; n = 11,438), Admixed American (AMR; n = 2,447), East Asian (EAS; n = 106,992), and European (EUR; n = 388,313) ancestry. SmkCes was defined by comparing current and former smokers, with positive effect estimates indicating greater genetic liability to smoking persistence.

#### 1.2 1000 Genomes Project Reference Panel

The 1KG Phase 3 reference panel includes 2,504 individuals from 26 populations spanning five continental ancestry groups: AFR, AMR, EAS, EUR, and South Asian (SAS).^15^ We used 1KG as the linkage disequilibrium (LD) reference for PGS construction, principal component (PC) analysis, genetic ancestry inference, and PGS calibration.

#### 1.3 All of Us Research Program Target Cohort

The National Institutes of Health (NIH) AoU Research Program is a large US cohort designed to advance precision medicine through diverse participant representation, with genomic, survey, and electronic health record data.^16,17^ For this study, we included participants with available genomic data who could be classified as currently or previously smoking based on survey responses (Supplementary Methods 1 and Figure S2). Related individuals were excluded using AoU-provided kinship information (kinship coefficient >0.1). The final cohort comprised 139,129 participants (58,872 current and 80,257 former smokers). Participants reporting a sex other than male or female (n = 1,529) were retained in descriptive summaries but excluded from statistical analyses.

#### 1.4 Smoking and Demographic Variables

Smoking variables included age at initiation, age at cessation, cigarettes smoked per day, and pack-years. Sociodemographic variables included age, sex, educational attainment, and household income. Education and income were modeled categorically, with approximate years of education and income midpoints derived for descriptive presentation in Table 1. Detailed variable definitions and survey response mappings are provided in Supplementary Methods 1.

**Table 1.** Cohort characteristics overall and by smoking status.

| Characteristic | Overall<br>(N = 139,129) | Smoking status |  |
| --- | --- | --- | --- |
|  |  | Current<br>(n = 58,872) | Former<br>(n = 80,257) |
| **Age at survey, mean (SD), years** | 54.6 (15.6) | 48.0 (13.5) | 59.5 (15.3) |
| **Sex at birth, No. (%)** |  |  |  |
| Male | 66,042 (47.5) | 30,699 (52.1) | 35,343 (44.0) |
| Female | 71,558 (51.4) | 27,260 (46.3) | 44,298 (55.2) |
| Other or missing | 1,529 (1.1) | 913 (1.6) | 616 (0.8) |
| **Race and ethnicity, No. (%)** |  |  |  |
| White | 78,287 (56.3) | 21,791 (37.0) | 56,496 (70.4) |
| Black | 28,023 (20.1) | 21,160 (35.9) | 6,863 (8.6) |
| Asian | 1,732 (1.3) | 518 (0.9) | 1,214 (1.5) |
| Hispanic | 14,783 (10.6) | 7,231 (12.3) | 7,552 (9.4) |
| Multiracial | 9,739 (7.0) | 4,612 (7.8) | 5,127 (6.4) |
| Other or missing | 6,565 (4.7) | 3,560 (6.0) | 3,005 (3.7) |
| **Years of education, mean (SD), years** | 14.0 (2.8) | 12.7 (2.5) | 14.8 (2.6) |
| **Household income, mean (SD), \$1000** | 64.1 (63.1) | 39.8 (51.9) | 82.0 (64.5) |
| **Age at smoking initiation, years** |  |  |  |
| No. (%) with data | 129,188 (92.9) | 51,636 (87.7) | 77,552 (96.6) |
| Mean (SD) | 17.9 (6.5) | 18.2 (7.8) | 17.7 (5.5) |
| **Smoking duration, years** |  |  |  |
| No. (%) with data | 128,417 (92.3) | 51,636 (87.7) | 76,781 (95.7) |
| Mean (SD) | 23.2 (14.9) | 29.7 (14.9) | 18.8 (13.3) |
| **Cigarettes smoked per day** |  |  |  |
| No. (%) with data | 125,524 (90.2) | 49,043 (83.3) | 76,481 (95.3) |
| Mean (SD) | 13.1 (11.4) | 12.3 (9.8) | 13.6 (12.2) |
| **Pack-years of smoking** |  |  |  |
| No. (%) with data | 119,122 (85.6) | 45,604 (77.5) | 73,518 (91.6) |
| Mean (SD) | 16.9 (20.1) | 19.9 (20.5) | 15.3 (19.6) |

### 2 Polygenic Score (PGS) Construction

Smoking cessation PGSs were constructed using PRS-CSx, a Bayesian polygenic prediction method that jointly models ancestry-specific GWAS summary statistics while accounting for population differences in LD.^18^ GSCAN2 SmkCes summary statistics from AFR, AMR, EAS, and EUR populations were jointly modeled using the corresponding ancestry-matched 1KG LD reference panels, with variants restricted to HapMap3 SNPs shared across discovery, reference, and target datasets. The PRS-CSx meta-analysis option (--meta=True) combined ancestry-specific posterior effect estimates through inverse-variance weighting, yielding weights for 1,291,355 SNPs. PGSs were calculated in AoU and 1KG using PLINK 2.0 (--score),^19^ with 1KG scores generated for use in subsequent calibration of AoU score.

### 3 Genetic Population Structure and PGS Calibration

#### 3.1 Genetic Principal Component Analysis

PCA was performed in 1KG, with AoU participants projected onto the resulting PC space. Analyses used shared HapMap3 variants following quality control and LD pruning, retaining 86,003 variants (Supplementary Methods 2 and Figure S3).^20^ The first five PCs were used for genetic ancestry inference and PGS calibration, while the first ten PCs were included in association models.

#### 3.2 Genetic Ancestry Inference

Genetic ancestry was inferred using a random forest classifier trained on the first five PCs of 1KG participants and their ancestry labels. AoU participants were assigned to the group with the highest predicted probability when this probability was at least 80%, otherwise, they were classified as “Other.” These labels represent reference-based groupings of continuous genetic variation and are used for illustrative and analytical purposes. They should not be interpreted as discrete biological populations or as equivalent to self-identified race or ethnicity.

#### 3.2 PGS Calibration

To reduce differences in the PGS distribution associated with genetic population structure, we applied a previously described two-step regression-based calibration procedure.^21,22^ Calibration equations and implementation details are provided in Supplementary Methods 3, with score distributions before and after calibration shown in Supplementary Figure S4. The calibrated scores were subsequently standardized within AoU to a mean of zero and standard deviation of one.

### 4 Statistical Analyses

Participants were divided into PGS deciles to examine the proportion of current smokers across the score distribution, then grouped into the bottom 10%, middle 80%, and top 10% for subsequent analyses.

Smoking cessation timing was examined using survival analysis, with age as the time scale. Participants entered the risk set at their reported age of smoking initiation. Former smokers experienced the event at their reported age of smoking cessation, while current smokers were right censored at their age at assessment. Kaplan-Meier (KM) analysis was used to construct unadjusted cumulative cessation curves with 95% confidence intervals (CIs) across the PGS groups. Covariate-adjusted cumulative cessation curves were estimated through marginal standardization of a fitted Cox proportional hazards model,^23^ averaging model-based cessation probabilities across participants’ observed covariate profiles for each PGS group. The Cox model adjusted for sex at birth, educational attainment, annual household income, cigarettes smoked per day, and the first ten genetic PCs. Both unadjusted and adjusted curves were used to estimate median cessation age, defined as the age at which cumulative cessation probability reached 50%, and cumulative cessation probabilities by ages 40, 50, and 60. For the marginally standardized estimates, 95% CIs were obtained using 200 bootstrap resamples.

Cox proportional hazards regression was also used to estimate hazard ratios (HRs) and 95% CIs for smoking cessation across PGS groups, using the bottom 10% as the reference. These analyses were repeated within genetically inferred ancestry groups to examine associations across diverse genetic backgrounds. The same PGS group thresholds and covariate adjustment strategy were applied across ancestry groups.

Additional analyses evaluated associations between continuous PGS and smoking cessation timing using Cox proportional hazards regression. Hazard ratios were reported per standard deviation increase in PGS, with adjustment for the previously described covariates.

All statistical analyses were conducted in RStudio (R version 4.3.1). Ethics Approval

The All of Us Research Program was approved by the All of Us Institutional Review Board, and all participants provided informed consent. This study used Controlled Tier data accessed through the All of Us Researcher Workbench under a Data Use and Registration Agreement with Washington University in St. Louis.

## Results

### Study Population and Polygenic Score Distribution

The study cohort included 139,129 AoU participants, and of these, 58,872 (42.3%) currently smoked and 80,257 (57.7%) formerly smoked. Participant characteristics according to smoking status are presented in Table 1. The cohort was diverse in self-reported race/ethnicity, with 56.3% of participants identifying as White. Participants who currently smoked were younger on average than participants who formerly smoked (48.0 vs. 59.5 years) and included a higher proportion of males (52.1% vs. 46.0%). They also had fewer average years of education (12.7 vs. 14.8 years), lower mean household income ($39,800 vs. $82,000), and greater smoking exposure, including longer smoking duration (29.7 vs. 18.8 years) and higher pack-years (19.9 vs. 15.3), despite slightly lower average cigarettes smoked per day (12.3 vs. 13.6).

The multi-ancestry smoking cessation PGS was constructed using GSCAN2 summary statistics and PRS-CSx, followed by ancestry-informed calibration using the 1KG reference panel (Supplementary Methods 3). Calibration reduced ancestry-associated differences in the PGS distribution, resulting in more comparable score distributions across genetic ancestry groups (Supplementary Figure S4).

### Smoking Persistence Increases Across the Polygenic Score Distribution

We examined smoking persistence across the PGS distribution by dividing participants into deciles. The proportion of current smokers increased across successive PGS deciles (Figure 1A, Supplementary Table S1), indicating greater smoking persistence among participants with higher PGS.

**Figure 1.**
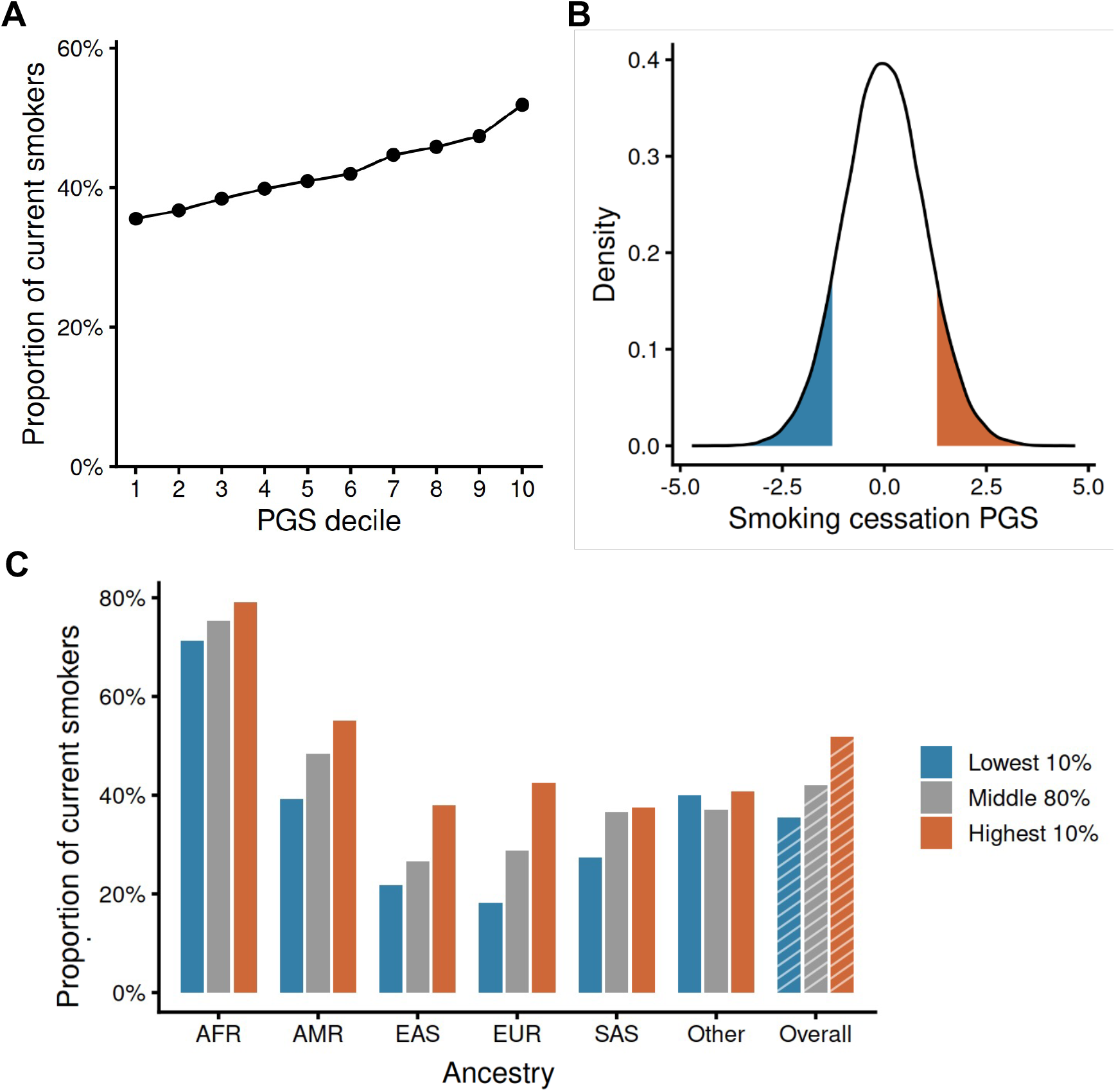
Smoking cessation patterns across the polygenic score distribution. (A) Proportion of current smokers across polygenic score (PGS) deciles. (B) Distribution of standardized PGS values, highlighting the bottom 10% and top 10%. (C) Proportion of current smokers across PGS groups, overall and by genetically inferred ancestry group. PGS values were derived from a multi-ancestry genome-wide association study of smoking cessation, with higher scores indicating greater genetic propensity for smoking persistence.

To further characterize differences at the extremes of the distribution, participants were categorized into the bottom 10%, middle 80%, and top 10% of PGS values (Figure 1B). The proportion of current smokers was 35.5%, 42.0%, and 51.9%, respectively (Figure 1C). Within genetic ancestry groups, the proportion of current smokers generally increased with higher PGS, although the overall unadjusted prevalence of current smoking varied substantially across ancestry groups. Current smoking was most prevalent in the AFR group (75.2%) and least prevalent in the EAS (27.3%) and EUR (29.0%) groups (Supplementary Table S2).

### Individuals with Higher Polygenic Scores Experience Later Smoking Cessation

We subsequently examined whether differences across the PGS distribution translated into differences in cessation timing. Unadjusted KM curves showed that participants with higher PGS had lower cumulative probabilities of smoking cessation across age (Supplementary Figure S5). These differences were also reflected in the median age at smoking cessation, defined as the age at which the cumulative probability of cessation reached 50%. KM-estimated median cessation ages were 38 (95% CI: 37-39), 44 (95% CI: 43-44), and 50 (95% CI: 50-51) years for participants in the bottom 10%, middle 80%, and top 10%, respectively (Table 2), representing a 12-year difference between the extremes of the PGS distribution. Cumulative cessation probabilities at ages 40, 50, and 60 followed a similar pattern, with lower probabilities among participants with higher PGS.

**Table 2.**
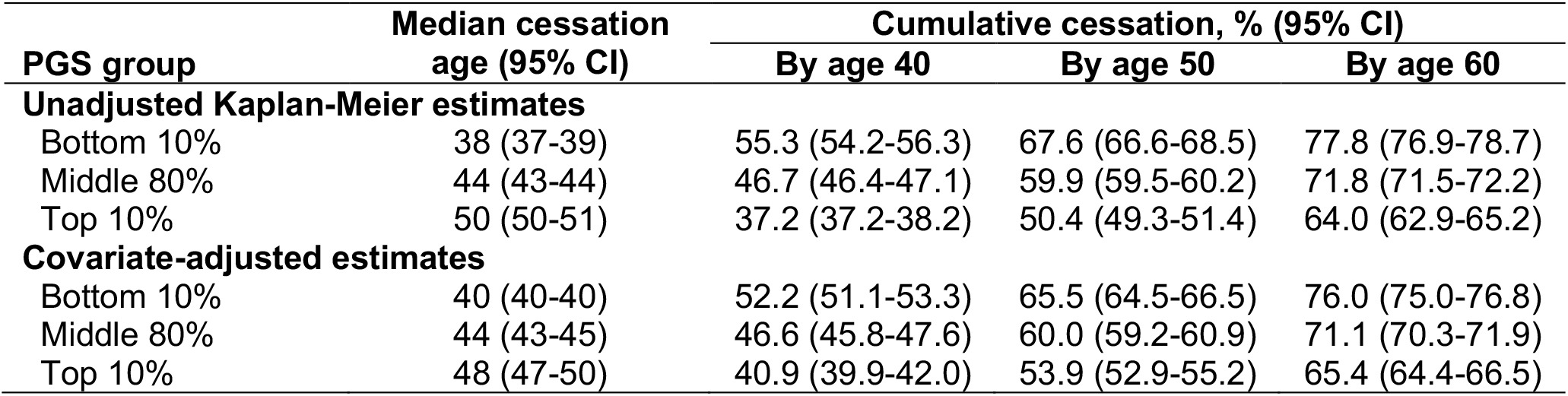
Median cessation age and cumulative cessation by PGS group.

| PGS group | Median cessation age (95% CI) | Cumulative cessation, % (95% CI) |  |  |
| --- | --- | --- | --- | --- |
|  |  | By age 40 | By age 50 | By age 60 |
| Unadjusted Kaplan-Meier estimates |  |  |  |  |
| Bottom 10% | 38 (37-39) | 55.3 (54.2-56.3) | 67.6 (66.6-68.5) | 77.8 (76.9-78.7) |
| Middle 80% | 44 (43-44) | 46.7 (46.4-47.1) | 59.9 (59.5-60.2) | 71.8 (71.5-72.2) |
| Top 10% | 50 (50-51) | 37.2 (37.2-38.2) | 50.4 (49.3-51.4) | 64.0 (62.9-65.2) |
| Covariate-adjusted estimates |  |  |  |  |
| Bottom 10% | 40 (40-40) | 52.2 (51.1-53.3) | 65.5 (64.5-66.5) | 76.0 (75.0-76.8) |
| Middle 80% | 44 (43-45) | 46.6 (45.8-47.6) | 60.0 (59.2-60.9) | 71.1 (70.3-71.9) |
| Top 10% | 48 (47-50) | 40.9 (39.9-42.0) | 53.9 (52.9-55.2) | 65.4 (64.4-66.5) |

These differences persisted following adjustment for sex, educational, income, cigarettes smoked per day, and the first ten genetic PCs. Covariate-adjusted cumulative cessation curves derived through marginal standardization of the Cox proportional hazards model continued to show lower cessation probabilities among participants with higher PGS (Figure 2). The adjusted median age at cessation was 40 years (95% CI: 40-41) in the bottom 10%, compared with 44 years (95% CI: 43-45) in the middle 80% and 48 years (95% CI: 47-50) in the top 10% (Table 2). Although attenuated following covariate adjustment, the estimated median cessation ages correspond to an 8-year difference between the bottom and top deciles, compared with 12 years based on the unadjusted KM estimates. By age 40, the adjusted cumulative probability of cessation was 52.2% in the bottom 10%, compared with 40.9% in the top 10%. Cumulative cessation probabilities increased with age across all PGS groups, while remaining consistently lower among participants with higher PGS at ages 50 and 60 (Table 2).

**Figure 2.**
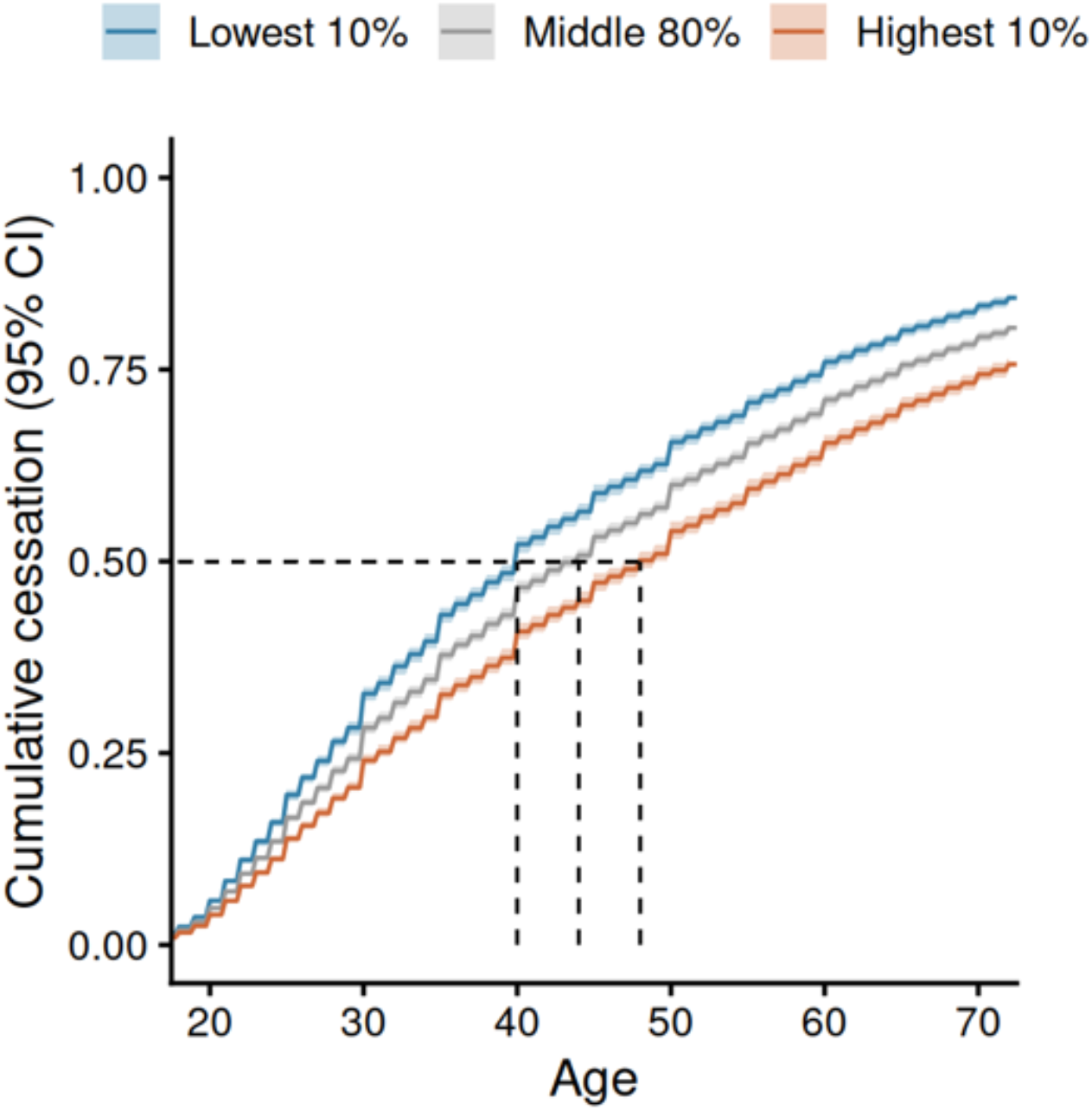
Adjusted cumulative probability of smoking cessation by polygenic score group. Cumulative probabilities of smoking cessation across the bottom 10%, middle 80%, and top 10% of the PGS distribution. Curves were estimated using marginal standardization of Cox proportional hazards models adjusted for sex at birth, educational attainment, household income, cigarettes smoked per day, and the first 10 genetic principal components.

### Polygenic Scores Are Associated with Smoking Cessation Across Genetic Ancestries

Cox proportional hazards regression further quantified associations between PGS groups and smoking cessation (Figure 3). Compared with participants in the bottom 10% of the PGS distribution, those in the middle 80% had a 17% lower hazard of smoking cessation (HR = 0.83, 95% CI: 0.81-0.85), while those in the top 10% had a 33% lower hazard (HR = 0.67, 95% CI: 0.65-0.70) (Figure 3, Supplementary Table S3). A similar pattern was observed across genetic ancestry groups, with participants in the top 10% generally exhibiting lower cessation hazards than those in the middle 80%, relative to the bottom 10%. Effect estimates varied in magnitude and precision, with wider confidence intervals observed in the EAS (n = 1,050) and SAS (n = 420) groups, which had the smallest sample sizes.

**Figure 3.**
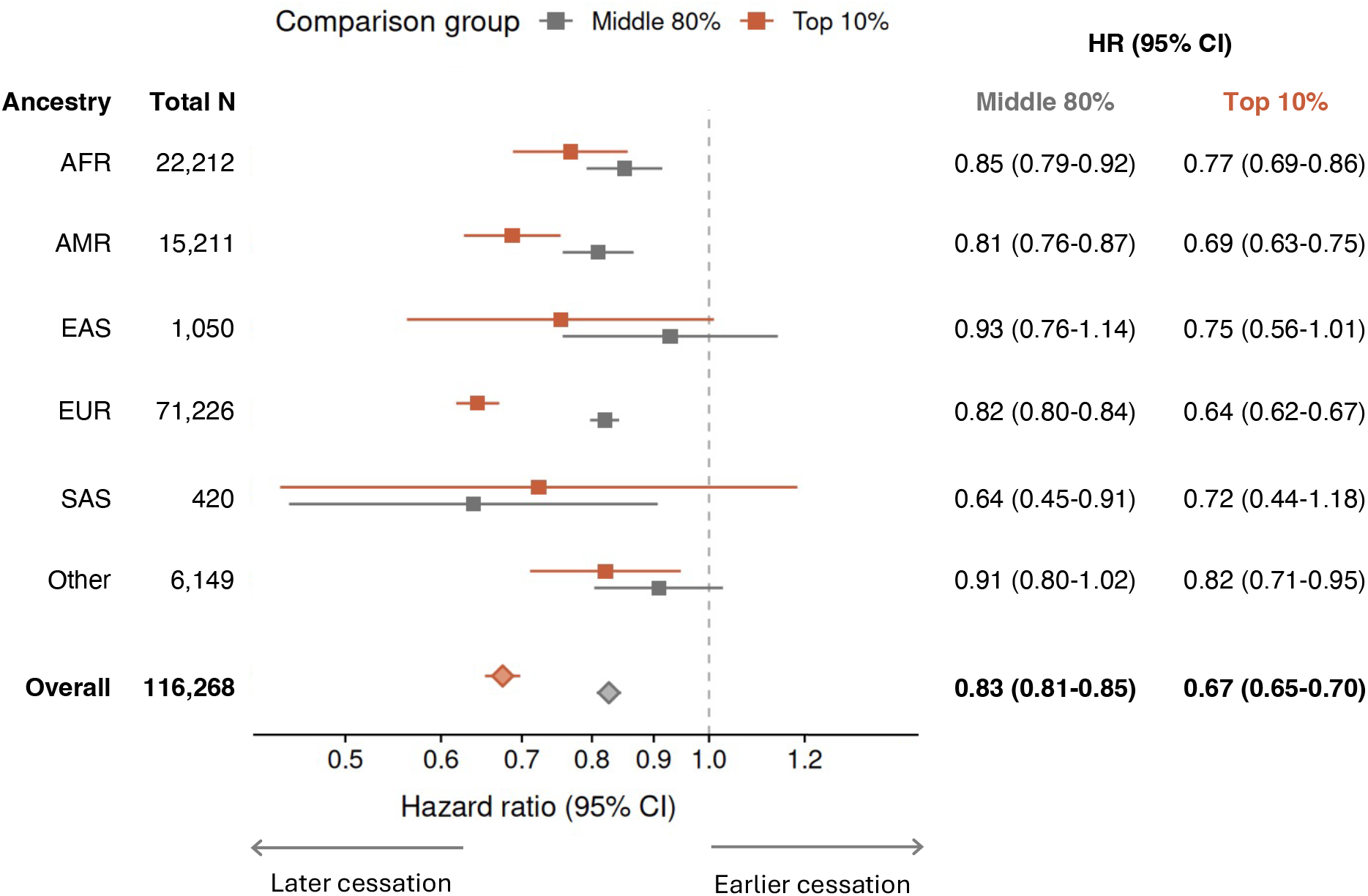
Association of polygenic score groups with smoking cessation across genetic ancestry groups. Adjusted hazard ratios (HRs) and 95% CIs for smoking cessation comparing the middle 80% and top 10% of the PGS distribution with the bottom 10% (reference), overall and by genetically inferred ancestry group. Estimates were obtained from Cox proportional hazards models adjusting for sex at birth, educational attainment, household income, cigarettes smoked per day, and the first 10 genetic principal components. HRs below 1 indicate a lower hazard of smoking cessation, whereas HRs above 1 indicate a higher hazard relative to the reference group. The dashed vertical line indicates an HR of 1.

To complement the group-based analyses, we examined associations across the continuous PGS distribution. Higher PGS was associated with a lower hazard of smoking cessation (HR = 0.89, 95% CI: 0.88-0.90), consistent with the smoking cessation patterns observed across PGS groups (Supplementary Figure S6).

## Discussion

The PGS was associated with the timing of smoking cessation. Individuals in the highest decile of our smoking cessation PGS had an estimated median cessation age 8 years later than those in the lowest decile, even after adjustment for genetic ancestry, socioeconomic factors, and smoking intensity. Individuals with higher scores were more likely to quit smoking later, with similar patterns of association observed across genetic ancestry groups. These findings suggest that a PGS derived from a GWAS of current versus former smoking captures clinically meaningful differences not only in smoking persistence but also in the timing of cessation.

Our findings extend previous research linking genetic variation to smoking cessation outcomes. An earlier study of the CHRNA5 variant rs16969968 identified a 4-year difference in median cessation age between high- and low-risk genotypes.^24^ Subsequent studies by Bray et al. identified associations between smoking-related PGSs and cessation outcomes, although results varied across scores and genetic ancestry groups.^25,26^ More recently, Chen et al. developed a 175-variant PGS for difficulty quitting smoking and found associations with cessation in UK Biobank (UKB) and AoU across genetic ancestry groups.^22^ Building on these findings, our genome-wide PGS incorporating more than one million variants demonstrated greater differentiation in cessation outcomes between individuals at the extremes of genetic risk, including an approximately 8-year difference in adjusted median cessation age.

The timing of smoking cessation is particularly important, as large epidemiological studies have shown that quitting reduces mortality at any age, with greater benefits associated with earlier cessation.^4,5,27^ In our study, the difference in estimated median cessation age between the highest and lowest PGS deciles was 12 years in unadjusted analyses and 8 years following adjustment for genetic population structure, socioeconomic characteristics, and smoking intensity. To contextualize this difference, eight additional years of smoking at the average intensity of 12 cigarettes per day reported by current smokers in our cohort would correspond to approximately 35,000 additional cigarettes smoked, or 4.8 additional pack-years of tobacco exposure. The increased exposure to cigarette smoking elevates the risk of developing cardiovascular diseases, chronic obstructive pulmonary disease (COPD), and cancer.^28^ The persistence of this 8-year difference after covariate adjustment is particularly notable, given the broader socioeconomic context of smoking cessation, in which education, income, and healthcare access influence the opportunities and resources available for quitting.^29,30^

The association between PGS and cessation timing also raises the possibility of identifying individuals at increased risk of delayed smoking cessation earlier in their smoking history. Despite the availability of effective smoking cessation treatments, their use remains limited. In a nationally representative study of US adults, VanFrank et al. reported that only 38.3% of adults who had recently attempted or successfully quit smoking used counseling or medication, with 36.3% using cessation medication.^31^ Earlier identification of individuals at greater risk of persistent smoking could potentially inform more aggressive treatment for smoking cessation, including earlier and more intensive behavioral and pharmacological interventions.

An important consideration is the generalizability of these associations across genetically diverse populations. GWASs have historically been dominated by European-ancestry samples, contributing to reduced portability of PGSs across populations.^32^ Saunders et al. documented this limitation in substance use-related PGSs, including smoking cessation, finding that predictive accuracy declined with increasing genetic distance from the discovery population.^33^ This is consistent with similar patterns of reduced portability that have been observed across a broader range of complex traits.^34^ Although large multi-ancestry genetic studies have expanded the representation of diverse populations, substantial imbalances in discovery sample sizes remain, motivating approaches such as PRS-CSx that integrate ancestry-specific genetic association estimates and linkage disequilibrium information to improve cross-population prediction.^18^ Using this approach, we observed associations between higher PGS and later smoking cessation across genetic ancestry groups, although effect estimates were less precise in the smaller EAS and SAS samples. These findings suggest that the association between polygenic liability and cessation timing is not confined to a single genetic ancestry group, while emphasizing the need for larger and more balanced populations to evaluate its generalizability. Importantly, differences in smoking cessation patterns across populations may also reflect social, environmental, and structural influences, and should not be interpreted as genetically determined.^35,36^

Several limitations should be considered. First, smoking history was derived from participant-reported survey data and may be subject to recall or reporting error, particularly for retrospectively reported ages of smoking initiation and cessation, and quantity of cigarettes smoked. Second, the current-versus-former smoking phenotype does not capture repeated quit attempts, relapse, nicotine dependence, cessation treatment, or the circumstances surrounding quitting. Finally, unequal ancestry representation in discovery and target samples limited the precision of some ancestry-specific estimates. Differences in overall cessation rates, including higher rates among participants of African ancestry, may also contribute to variation in observed PGS associations across ancestry groups.

In conclusion, our findings demonstrate that polygenic liability to smoking persistence is associated with substantial differences in the timing of smoking cessation, with associations observed across genetically diverse populations. These findings highlight the potential of PGSs to identify individuals at increased risk of delayed cessation earlier in their smoking history, which could inform more timely and targeted cessation interventions to reduce prolonged tobacco exposure and its associated health risks.

## Data Availability

All data produced in the present work are contained in the manuscript

## Funding and Acknowledgements

This work was supported by the National Institutes of Health (NIH) National Cancer Institute (NCI) grant numbers U19CA203654 (A.C., A.T.R., L.J.B. and Y.C.), R01CA268030 (A.T.R.), UG3CA305777 (A.T.R.) and P30CA091842 (A.T.R.); National Institute on Alcohol Abuse and Alcoholism (NIAAA) grant numbers U10AA008401 (A.C. and L.J.B.) and R01AA031968 (L.J.B.); National Institute on Drug Abuse (NIDA) grant numbers K02DA065836 (A.T.R.), R01DA056050 (A.T.R.), K01DA057376 (G.S.), R01DA065287 (G.S.) and P50 DA054071 (E.O.J.); National Center for Advancing Translational Sciences (NCATS) grant number UL1TR002345 (A.T.R.) and the Taylor Family Institute for Innovative Psychiatric Research. This research was also supported in part by the Intramural Research Program (IRP) of the NIH. The contributions of the NIH authors were made as part of their official duties as NIH federal employees, are in compliance with agency policy requirements, and are considered Works of the United States Government. However, the findings and conclusions presented in this paper are those of the authors and do not necessarily reflect the views of the NIH or the U.S. Department of Health and Human Services.

## AI Use Disclosure

ChatGPT (OpenAI, GPT-6) was used to assist with portions of manuscript language editing, as well as refining analytical code for consistency and conciseness. All AI-assisted content and code were reviewed and verified by the authors.

