## Supplemental Information for "A Polygenic Score for Smoking Persistence Captures Differences in Cessation Age"

*Chaloemtoem et al.*

### Table of Contents

**Supplementary Methods** ... 2

**Supplementary Methods 1.** Smoking phenotype and covariate definitions ... 2

**Supplementary Methods 2.** Genetic principal component analysis ... 3

**Supplementary Methods 3.** Two-step regression-based PGS calibration ... 4

**Supplementary Tables** ... 5

**Table S1.** Proportion of current smokers across PGS deciles ... 5

**Table S2.** Proportion of current smokers by ancestry and PGS group ... 6

**Table S3.** Adjusted HR for smoking cessation relative to the lowest decile ... 7

**Supplementary Figures** ... 8

**Figure S1.** Overview of the study design and analytical workflow ... 8

**Figure S2.** Defining current and previous smoking in AoU ... 9

**Figure S3.** Genetic principal component analysis of 1KG and AoU ... 10

**Figure S4.** PGS distributions before and after calibration ... 11

**Figure S5.** Kaplan-Meier estimated cumulative cessation across PGS groups ... 12

**Figure S6.** Association between continuous PGS and time to smoking cessation ... 13

**References** ... 14

#### Supplementary Methods

**Supplementary Methods 1.** Smoking phenotype and covariate definitions

Smoking variables were derived from All of Us (AoU) Participant Provided Information (PPI) survey data and identified using AoU PPI concept IDs.

Smoking status: Smoking status was defined using participants' responses to two questions (Supplementary Figure S1). First, participants were asking about their lifetime cigarette use (concept ID 1585857: Have you smoked at least 100 cigarettes in your entire life?). Participants who reported having smoked at least 100 cigarettes were then asked about their current smoking frequency (concept ID 1585860: Do you now smoke cigarettes every day, some days, or not at all?). Participants who responded currently smoking "Every day" or "Some days" were assigned current smoking status, while those responding "Not at all" were assigned previous smoking status.

Age at smoking initiation and cessation: Age at smoking initiation was derived from the participant-reported response to "How old were you when you first started regular cigarette smoking?" (concept ID 1585864). For participants who formerly smoked, age at smoking cessation was derived from the response to "If you have completely stopped smoking cigarettes, how old were you when you stopped?" (concept ID 1585870). For participants who currently smoked, age at survey was assigned as the endpoint age, representing their age at last observation rather than an observed cessation age.

Smoking duration: Smoking duration was calculated as the interval between age at smoking initiation and age at smoking cessation among participants who formerly smoked and between age at smoking initiation and age at assessment among participants who currently smoked.

Cigarettes per day: Cigarettes smoked per day, representing smoking intensity, was based on participants reported average number of cigarettes smoked per day over their smoking history (concept ID 1586162: On average, over the entire time that you smoked, how many cigarettes did you smoke each day?). For participants without this measure, reported current cigarettes smoked per day (concept ID 1586159: On average, how many cigarettes do you smoke per day now?) was used.

Pack years: Cumulative smoking exposure was estimated as pack-years, calculated as cigarettes smoked per day divided by 20 and multiplied by smoking duration in years.

Socioeconomic variables: Educational attainment and household income were obtained from the AoU Basics survey data (concept ID 1585940: What is the highest grade or year of school you completed? And concept ID 1585375: What is your annual household income from all sources?) Missing values for the original educational attainment (~2% of the data) and household income (~16% of the data) survey categories were imputed using multiple imputation by chained equations (MICE) with ordinal logistic regression. Following imputation, original educational attainment was consolidated into five categories and household income into four categories for use in analyses. For descriptive presentation in Table 1, educational attainment categories were assigned approximate years of education based on the typical duration associated with each educational level, while household income categories were assigned the midpoint of each reported income range. These derived continuous measures were used only for descriptive purposes.

**Supplementary Methods 2.** Genetic principal component analysis

To characterize genetic population structure within a common reference framework, principal component analysis (PCA) was performed using the 1000 Genomes (1KG) Phase 3 data as an external reference panel, with AoU participants projected into the same PC space. Variants were restricted to HapMap3 variants shared between 1KG and AoU. HapMap3 variants had previously undergone population-specific quality control, including Hardy-Weinberg equilibrium filtering (HWE p-value $\geq 10^{-6}$) and variant call-rate filtering (>95%).[1] Variant missingness was additionally assessed within the 1KG reference dataset, with variants exceeding 2% missing genotype calls excluded.

Variants were then LD pruned using PLINK 2.0 with a 200-variant sliding window, 50-variant step size, and $r^2$ threshold of 0.1 (--indep-pairwise 200 50 0.1). Following LD pruning, 86,003 variants were retained for PCA. PCA was performed in 1KG to obtain genetic PCs and corresponding variant loadings, which were then applied to AoU genotype data to project AoU participants into the 1KG-defined PC space. PCA projections are shown in Supplementary Figure S2.

**Supplementary Methods 3.** Two-step regression-based PGS calibration

The raw PGS was standardized through a two-step calibration process that adjusts for both mean and variance differences in score distribution due to population structure.[2,3] Calibration parameters were estimated using the 1KG dataset and then applied to the AoU cohort.

*Step 1: Mean Adjustment*

To adjust for systematic differences in average PGS values attributable to ancestry, a linear regression model was fit in 1KG with the raw score ($PGS_{raw}$) as the outcome and the top five genetic PCs as predictors:

$$\hat{u} = \beta_0 + \beta_1 \cdot PC_1 + \cdots + \beta_5 \cdot PC_5$$

The model estimates the expected mean score ($\hat{u}$) associated with ancestry. Estimated coefficients from this model ($\beta$) were used to predict $\hat{u}$ for each AoU participant based on their projected PC values.

*Step 2: Variance Adjustment*

Residuals from the previous model ($PGS_{raw} - \hat{u}$) were squared to serve as a proxy for PGS variance. A second linear regression was performed in the 1KG data, modeling the squared residuals as a function of the same five PCs:

$$\hat{\sigma}^2 = \gamma_0 + \gamma_1 \cdot PC_1 + \cdots + \gamma_5 \cdot PC_5$$

This model estimates the expected variance ($\hat{\sigma}^2$) associated with ancestry. Similarly, the estimated coefficients ($\gamma$) were used to compute $\hat{\sigma}^2$ for each AoU participant based on their projected PC values.

*Final PGS Adjustment*

Final genetic PC-adjusted scores for each AoU participant were calculated by subtracting the raw score by the predicted mean and then dividing by the predicted standard deviation (square root of the predicted variance):

$$PGS_{adj} = \frac{PGS_{raw} - \hat{u}}{\hat{\sigma}}$$

This adjustment reduces ancestry-associated differences in the PGS distribution, placing scores on a more comparable scale across genetic backgrounds.

#### Supplementary Tables

**Table S1.** Proportion of current smokers across PGS deciles (N = 139,129)

| Decile | N | N Current | % Current |
|---|---|---|---|
| 1 | 13913 | 4945 | 35.5 |
| 2 | 13913 | 5108 | 36.7 |
| 3 | 13913 | 5341 | 38.4 |
| 4 | 13913 | 5541 | 39.8 |
| 5 | 13913 | 5695 | 40.9 |
| 6 | 13913 | 5842 | 42.0 |
| 7 | 13913 | 6215 | 44.7 |
| 8 | 13913 | 6377 | 45.8 |
| 9 | 13913 | 6591 | 47.4 |
| 10 | 13912 | 7217 | 51.9 |

**Table S2.** Proportion of current smokers by ancestry and PGS group (N = 139,129)

| | Entire Sample | | | % Current by PGS group | | |
|---|---|---|---|---|---|---|
| **Ancestry** | **N** | **N Current** | **% Current** | **Bottom 10%** | **Middle 80%** | **Top 10%** |
| AFR | 30719 | 23108 | 75.2 | 71.2 | 75.4 | 79.1 |
| AMR | 18919 | 9151 | 48.4 | 39.2 | 48.4 | 55.2 |
| EAS | 1213 | 332 | 27.4 | 21.8 | 26.6 | 37.9 |
| EUR | 80459 | 23339 | 29.0 | 18.2 | 28.8 | 42.4 |
| SAS | 495 | 176 | 35.6 | 27.4 | 36.6 | 37.5 |
| Other | 7324 | 2766 | 37.8 | 39.9 | 37.0 | 40.8 |
| Overall[a] | 139129 | 58872 | 42.3 | 35.5 | 42.0 | 51.9 |

[a] Includes all participants across ancestry groups; not a separate ancestry group.

**Note:** Only percentages are presented by PGS group to maintain consistency and comply with the All of Us Research Program Data and Statistics Dissemination Policy,[4] as some cell counts were 20 or fewer.

**Table S3.** Adjusted HR for smoking cessation relative to the lowest decile

| Ancestry | Comparison | OR | CI lower | CI upper | P |
|---|---|---|---|---|---|
| AFR | Middle 80% vs Bottom 10% | 0.85 | 0.79 | 0.92 | 1.35e-05 |
| | Top 10% vs Bottom 10% | 0.77 | 0.69 | 0.86 | 2.34e-06 |
| AMR | Middle 80% vs Bottom 10% | 0.81 | 0.76 | 0.87 | 9.47e-10 |
| | Top 10% vs Bottom 10% | 0.69 | 0.63 | 0.75 | 1.73e-15 |
| EAS | Middle 80% vs Bottom 10% | 0.93 | 0.76 | 1.14 | 4.80e-01 |
| | Top 10% vs Bottom 10% | 0.75 | 0.56 | 1.01 | 5.79e-02 |
| EUR | Middle 80% vs Bottom 10% | 0.82 | 0.80 | 0.84 | 5.52e-45 |
| | Top 10% vs Bottom 10% | 0.64 | 0.62 | 0.67 | 1.98e-98 |
| SAS | Middle 80% vs Bottom 10% | 0.64 | 0.45 | 0.91 | 1.22e-02 |
| | Top 10% vs Bottom 10% | 0.72 | 0.44 | 1.18 | 1.97e-01 |
| Other | Middle 80% vs Bottom 10% | 0.91 | 0.80 | 1.02 | 1.15e-01 |
| | Top 10% vs Bottom 10% | 0.82 | 0.71 | 0.95 | 6.42e-03 |
| Overall[a] | Middle 80% vs Bottom 10% | 0.83 | 0.81 | 0.85 | 2.57e-57 |
| | Top 10% vs Bottom 10% | 0.67 | 0.65 | 0.70 | 1.08e-115 |

[a] Includes all participants across ancestry groups; not a separate ancestry group.

### Supplementary Figures

**Figure S1.** Overview of the study design and analytical workflow

Ancestry-specific smoking cessation genome-wide association study (GWAS) summary statistics from GSCAN2 were integrated with 1000 Genomes Project (1KG) reference data and All of Us Research Program (AoU) genotypes to construct a multi-ancestry polygenic score (PGS) using PRS-CSx. Variant effect estimates were combined across ancestry groups and applied to individual-level genotypes to calculate PGSs in 1KG and AoU. Using genetic principal components (PCs), calibration models developed in 1KG were applied to AoU to reduce ancestry-associated differences in PGS distributions. The calibrated PGS was then evaluated for associations with smoking cessation and for patterns across the PGS distribution.

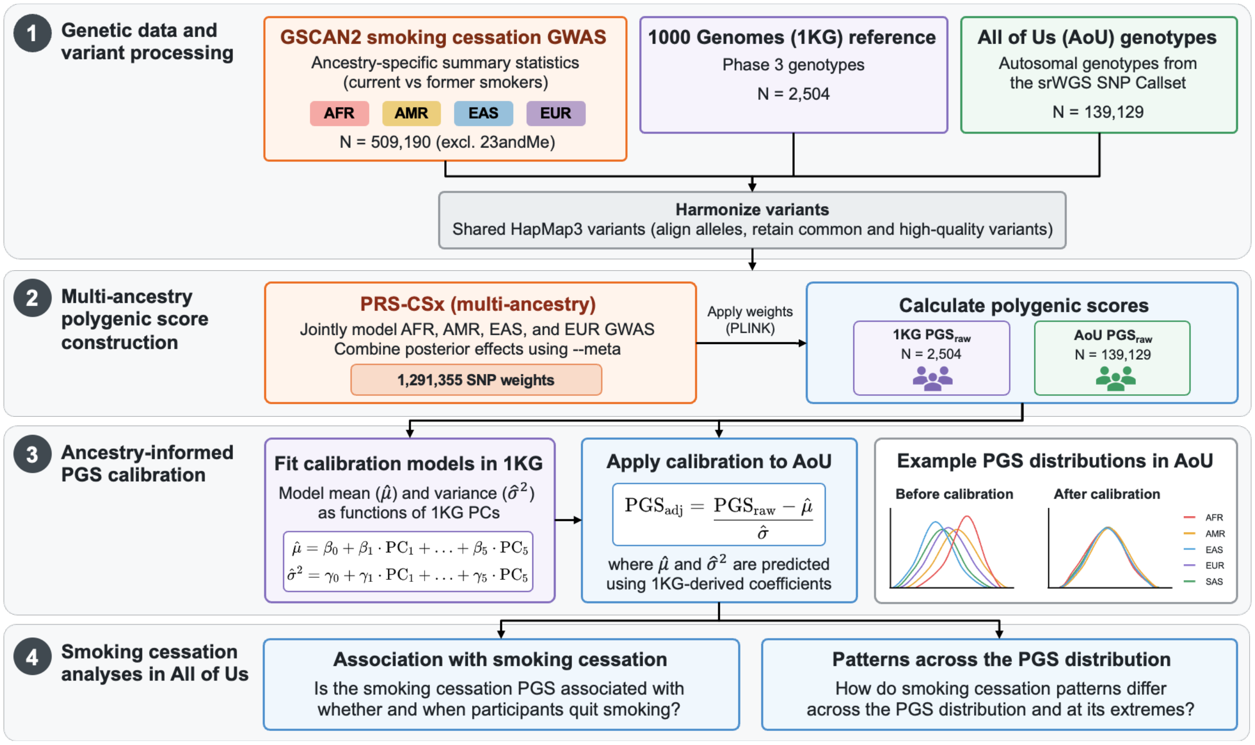

**Figure S2.** Defining current and previous smoking in AoU

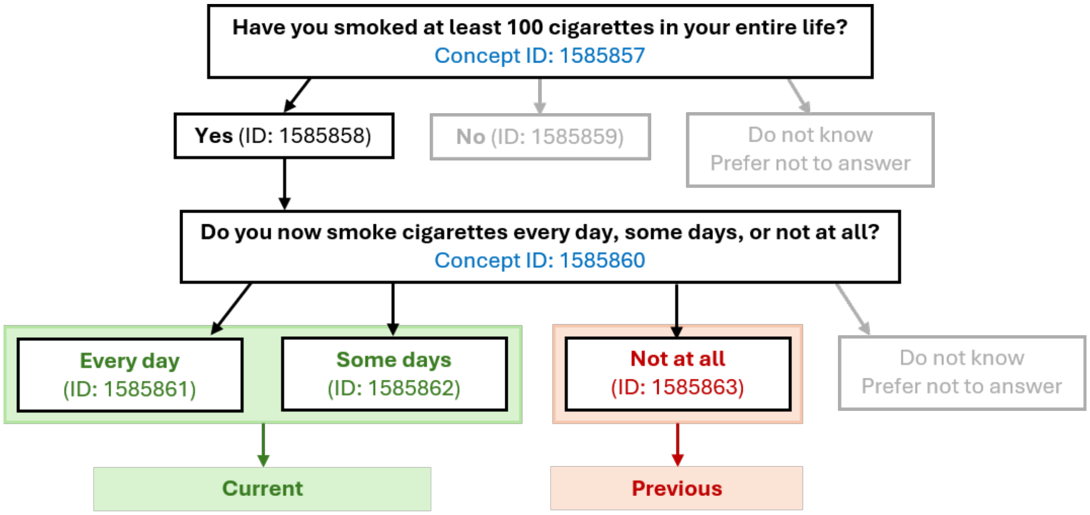

**Figure S3.** Genetic principal component analysis of 1KG and AoU

Principal component plots of **(A)** 1KG (N = 2,504) and **(B)** AoU (N = 139,129) participants, colored by genetically inferred ancestry.

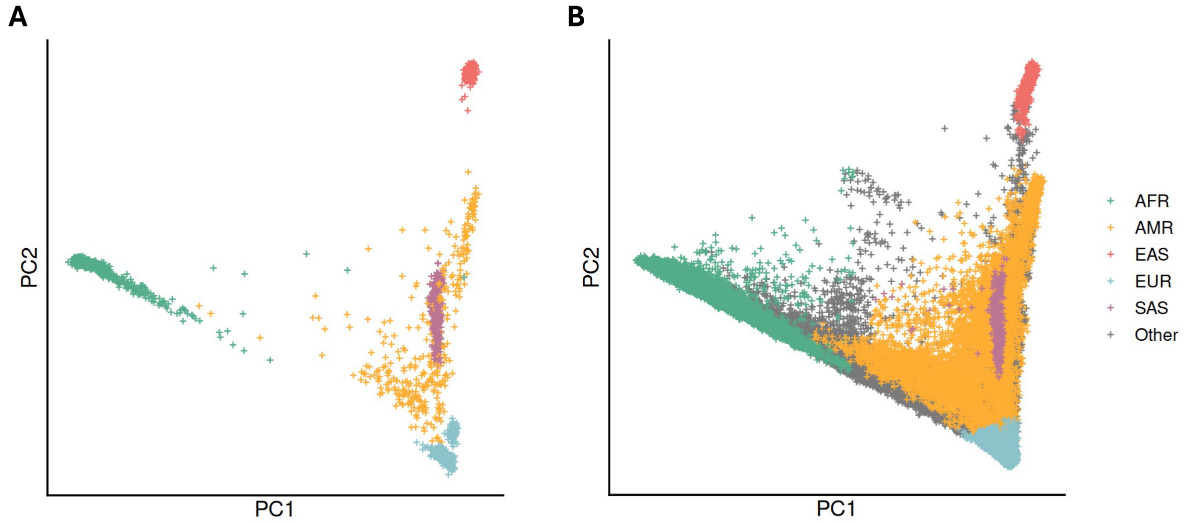

**Figure S4.** PGS distributions before and after calibration

Distributions of PGS scores in **(A)** 1KG (N = 2,504) and **(B)** AoU (N = 139,129) before and after calibration, colored by genetically inferred ancestry groups.

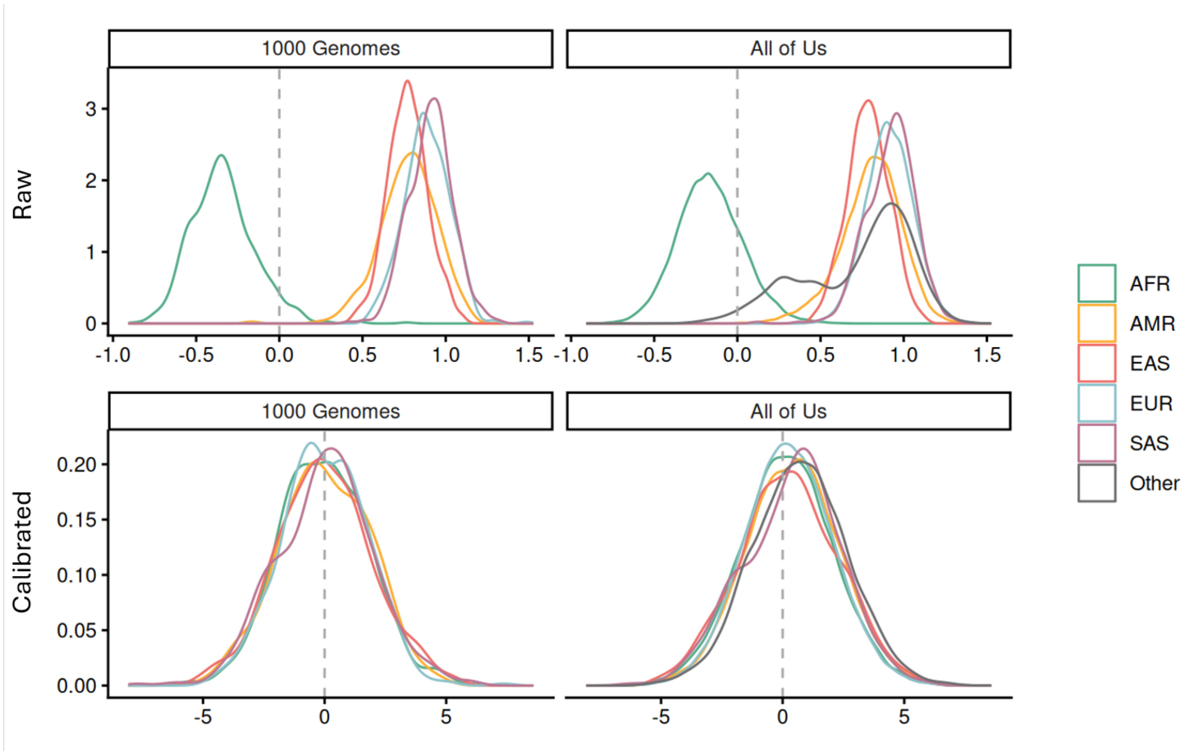

**Figure S5.** Kaplan-Meier estimated cumulative cessation across PGS groups

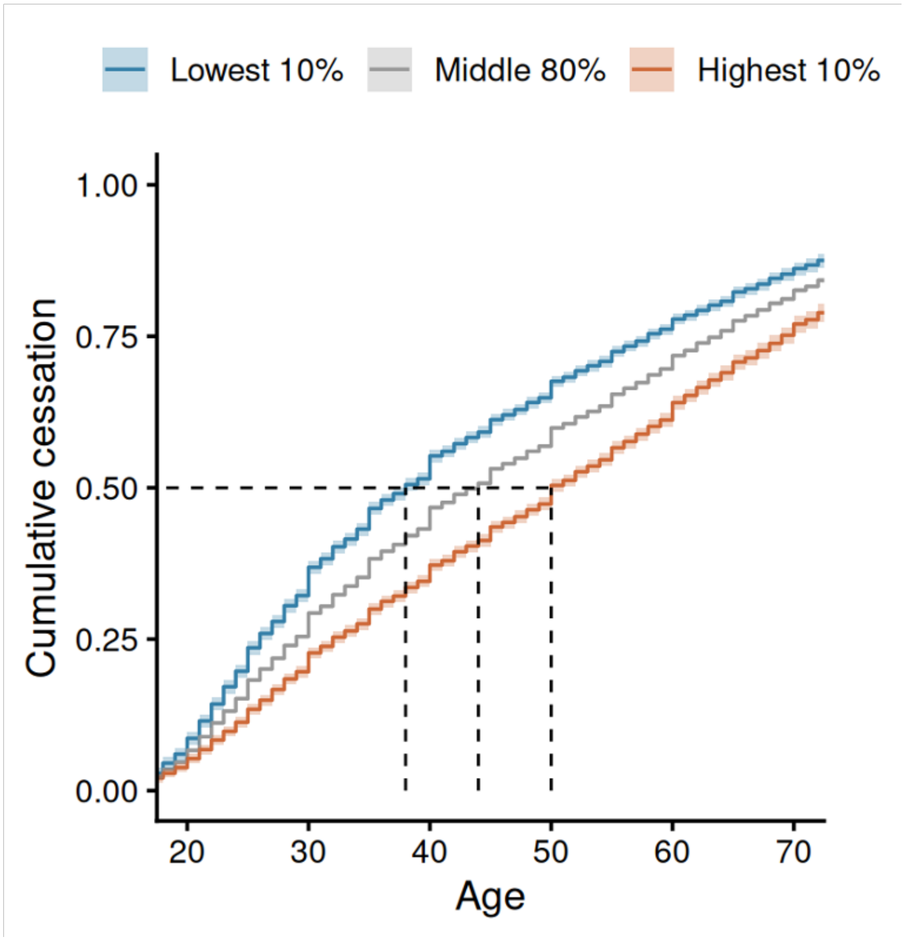

**Figure S6.** Association between continuous PGS and time to smoking cessation

Adjusted hazard ratios (HRs) for time to smoking cessation per 1-SD increase in PGS, overall and stratified by genetic ancestry. Squares represent ancestry-specific estimates, diamonds represent overall estimates not stratified by ancestry, and horizontal lines indicate 95% CIs. The dashed vertical line indicates the null association (HR = 1).

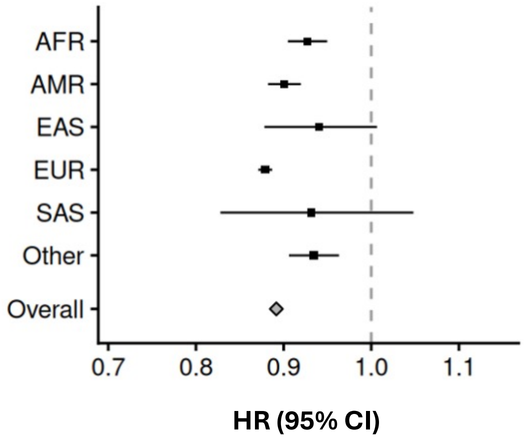


| N | % Quit | OR | 95% CI | P |
|---|---|---|---|---|
| 22212 | 28.7 | 0.93 | [0.91–0.95] | $8.46e^{-10}$ |
| 15211 | 56.0 | 0.90 | [0.88–0.92] | $6.55e^{-23}$ |
| 1050 | 76.2 | 0.94 | [0.88–1.01] | $7.63e^{-02}$ |
| 71226 | 73.1 | 0.88 | [0.87–0.89] | $7.88e^{-171}$ |
| 420 | 67.6 | 0.93 | [0.83–1.05] | $2.38e^{-01}$ |
| 6149 | 66.2 | 0.93 | [0.91–0.96] | $1.29e^{-05}$ |
| 116268 | 62.0 | 0.89 | [0.88–0.90] | $1.14e^{-196}$ |
